# Estimating the Failure Risk of Quarantine Systems for Preventing COVID-19 Outbreaks in Australia and New Zealand

**DOI:** 10.1101/2021.02.17.21251946

**Authors:** Leah Grout, Ameera Katar, Driss Ait Ouakrim, Jennifer A. Summers, Amanda Kvalsvig, Michael G. Baker, Tony Blakely, Nick Wilson

## Abstract

**Objectives:** To identify COVID-19 outbreaks and border control failures associated with quarantine systems in Australia and New Zealand and to estimate the failure risks.

**Design, setting, participants:** Observational epidemiological study of travellers transiting quarantine in Australia and New Zealand up to 15 June 2021.

**Main outcome measures:** The incidence of COVID-19 related failures arising from quarantine, and the failure risk for those transiting quarantine, estimated both per 100,000 travellers and per 1000 SARS-CoV-2 positive cases.

**Results:** Australia and New Zealand had 32 COVID-19 related failures arising from quarantine systems up to 15 June 2021 (22 and 10, respectively). One resultant outbreak involved an estimated 800 deaths and quarantine failures instigated nine lockdowns. The failure risk for those transiting quarantine was estimated at 5.0 failures per 100,000 travellers and 6.1 failures (95%CI: 4.0 to 8.3) per 1000 SARS-CoV-2 positive cases. The latter risk was two-fold higher in New Zealand compared with Australia. The full vaccination of frontline border workers could likely have prevented a number of quarantine system failures.

**Conclusions:** Quarantine system failures can be costly in terms of lives and economic impacts such as lockdowns. Ongoing improvements or alternatives to hotel-based quarantine are required.

## Introduction

New Zealand and Australian states have successfully eliminated community transmission of the pandemic virus ‘severe acute respiratory syndrome coronavirus 2’ (SARS-CoV-2),^1^ albeit with occasional outbreaks from imported cases that have typically been quickly brought under control. These two countries have primarily used hotel-based quarantine for citizens returning to their countries during the pandemic period, with 14 days of quarantine combined with polymerase chain reaction (PCR) testing and mask use in any shared spaces (eg, common exercise areas used in New Zealand, but not in most Australian states).

Converting hotels for quarantine purposes has the advantage of making use of a resource that would otherwise be underused during a pandemic, given declines in international tourism. However, the major disadvantage of hotel-based quarantine is that it is likely to be less effective than purpose-built quarantine facilities owing to shared spaces and lack of safe ventilation (as per World Health Organization advice on air flow^2^). Moreover, the consequences of leakage of the virus out of quarantine (eg, through facility workers) may be more severe given higher population density in urban settings where the hotels are located. Given these issues, we aimed to estimate the failure risk of quarantine systems in New Zealand and Australia in terms of the spread of ‘coronavirus disease 2019’ (COVID-19) infection into the community.

As of 13 June 2021, the rolling 7-day average number of COVID-19 vaccine doses administrated per 100 people was 0.46 in Australia and 0.31 in New Zealand.^3^ However, this was counted as single vaccine doses and does not equal the total number of people vaccinated (eg, the Pfizer/BioNTech vaccine which is currently used in New Zealand requires two doses).^3^ The majority of border workers in Australia and New Zealand have been vaccinated (eg, in New Zealand over 56,000 doses had been administered to border workers as of 28 March 2021,^4^ and all hotel quarantine workers in Victoria who have face-to-face contact with returned travellers received their first dose of the vaccine by the first week in April 2021^5^).

## Methods

We defined a quarantine system failure as where a border/health worker or person in the community with a link to the quarantine/isolation system, became infected with SARS-CoV-2. This definition included people infected in hospital from cases who had been transferred from a quarantine facility (as such cases were still in the 14-day quarantine process), but did not include pandemic virus transmission between returnees within the quarantine facilities.

We searched official websites in both countries, and for the eight states and territories in Australia, to identify outbreaks and border control failures associated with quarantine systems (searches conducted between 6 January and 23 June 2021). Where an outbreak source was uncertain (eg, the Auckland, New Zealand, August 2020 outbreak) we used the best available evidence to classify it as a quarantine failure or not. The decision to label an incident as a quarantine system failure was confirmed by all co-authors. We used two denominators: a) the estimated number of travellers who went through quarantine facilities during the 2020 year (data were for the period starting 1 April 2020 for Australia and 17 June 2020 for New Zealand) up to 15 June 2021; and b) the number of SARS-CoV-2 positive people who went through these facilities in this same time period. The unit of analyses were New Zealand, the eight Australian states and territories, and both countries combined.

For New Zealand, we used official data on both travellers going through the quarantine system^6^ along with official (Ministry of Health) data on SARS-CoV-2 positive cases,^7^ although there are some discrepancies in the information about when regular testing began in Managed Isolation and Quarantine (MIQ) facilities. For Australia we used overseas arrival data^8^ and health data.^9, 10^

## Results

The collated data for quarantine system failures are shown in Table 1, with specific details of each event in the Appendix (Table A1). In Australia, 22 failures were identified, one resultant outbreak caused over 800 deaths (Victoria’s second wave) and with eight lockdowns linked to quarantine system failures. In New Zealand, there were ten failures, with one causing an outbreak with three deaths, and also a lockdown.

**Table 1:**
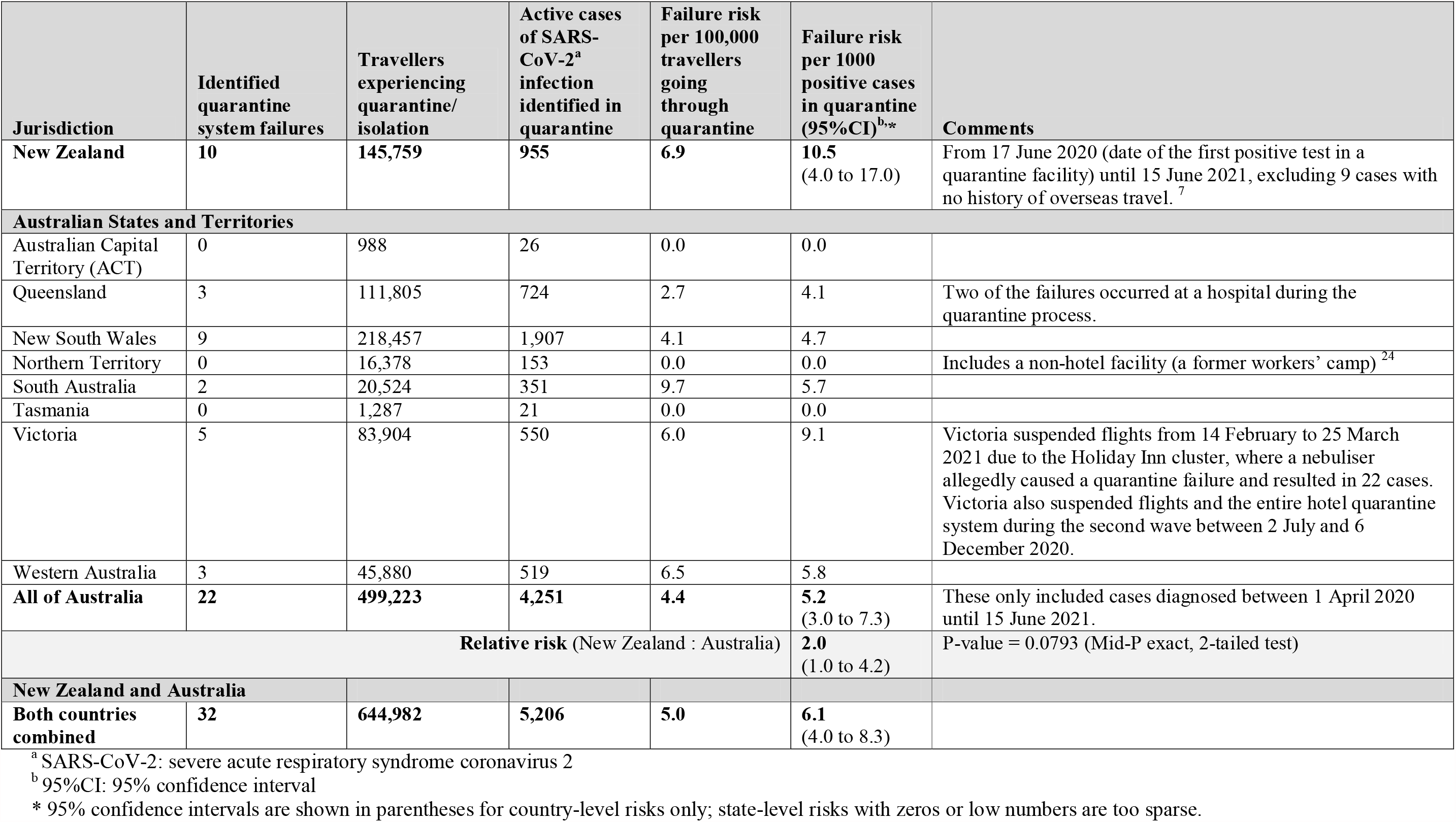
Identified quarantine system failures in Australia and New Zealand during the COVID-19 pandemic, the relevant denominator populations and estimated failure risks (with numerator and denominator data up to 15 June 2021)

Given our estimates of the number of travellers processed via quarantine systems (Table 1), the overall risks for both countries combined were one failure per 20,156 travellers, and one failure per 163 SARS-CoV-2 positive cases in quarantine. The combined data can also be interpreted as quarantine system failures leading to a lockdown response per 71,665 travellers; and approximately one death from COVID-19 per 803 travellers (using the 800 deaths estimate from Australia and the three deaths from New Zealand – although this figure is largely driven by the second wave in Victoria and is unlikely generalizable forward in time).

At the country level, there were 10.5 failures per 1000 SARS-CoV-2 positive cases transiting quarantine in New Zealand (95% confidence interval [CI]: 4.0 to 17.0), compared to 5.2 per 1000 SARS-CoV-2 positive cases in Australia (95%CI: 3.0 to 7.3) – a two-fold difference in risk (relative risk: 2.0, 95%CI: 1.0 to 4.2). The full vaccination of frontline border workers (Table A1) could likely have prevented a number of quarantine system failures due to the combined effects of vaccination lowering the risk of getting infected by 70% (as in the case of the Moderna vaccine^11^) or more (eg, the Pfizer/BioNTech vaccine is approximately 95% effective at preventing SARS-CoV-2 infection for wild-type and Beta variants for adults 16 years of age or older^13^), and the lesser duration of infectivity and lower peak infectivity for those infected.^12^

## Discussion

This analysis identified 32 failures of quarantine systems in Australia and New Zealand combined (up to 15 June 2021). The relatively higher failure risk per 1000 SARS-CoV-2 positive cases transiting quarantine in New Zealand versus Australia could reflect a lower quality approach in the former, with perhaps some of the difference due to greater detection in New Zealand from more border worker testing over a longer period.

These estimates are subject to chance variations due to the low numbers of failures. These estimates will also probably be an underestimate of all quarantine system failures, as not all of those infected will transmit the virus and start a detectable chain of transmission. Genomes of the first 649 viral isolates collected in New Zealand show that only 19% of virus introductions resulted in ongoing transmission of more than one additional case.^14^ Therefore, counts of border system failures are sensitive to how they are identified and defined. Indeed, with increased testing (eg, testing of people after leaving quarantine on day 16 as is now common in Australia), we may be detecting failures that previously would have been undetected.

Looking forward, the failure risks per month in New Zealand and Australia may increase, given that the proportion of travellers returning to these countries who are infected is increasing due to global intensification of the pandemic and the increasing infectivity of new SARS-CoV-2 variants.^15^ Indeed, there have been several clearly documented cases of spread *within* quarantine hotels (eg, two instances in Melbourne in February 2021, two instances in Sydney in April 2021, and one in South Australia in May 2021), highlighting the increased risk and evolving situation with more highly infectious variants arriving from overseas.

However, offsetting this trend will be measures such as the vaccination of quarantine workers. In New Zealand, the vaccination of border workers began in February 2021 with the Pfizer/BioNTech vaccine. However, vaccination does not fully protect against SARS-CoV-2 transmission, although a moderate degree of protection is likely. For example, infection rates were halved for the AstraZeneca vaccine,^16, 17^ reduced by 70% for the Moderna mRNA vaccine as indicated by using swab results for asymptomatic infection plus symptomatic cases,^11^ and reduced by 95% for the Pfizer/BioNTech mRNA vaccine as indicated by national surveillance data in Israel.^13^ For vaccinated people who are infected, primate study evidence suggests (consistent with expectation) that the infectivity is decreased in peak and duration,^12^ further protecting the border.

Furthermore, the level of testing of quarantine workers has been increasing (eg,^18^; which will find some failures before they have a chance to establish as an outbreak in the community). There have been other improvements in the quarantine systems over time (eg, improved security, introduction of mask wearing within quarantine settings, reduction in shared spaces, improved personal protective equipment (PPE) used by workers, and other procedures as detailed in both countries^19, 20^).

Another risk reduction practice would be using better or purpose-built facilities in rural locations as these have less risk from close contacts in central business district hotels and within-building spread from poor ventilation systems. To date, there have been no failures at the Howard Springs facility outside of Darwin, and the success of the facility was cited in the announcement in June 2021 of the construction of a new purpose-built quarantine facility in Victoria.^21^ Other infection prevention and control measures (eg, PPE) will remain important in all quarantine facilities.

Limitations of our analysis include residual uncertainty around the cause of some outbreaks (eg, the Auckland one in August 2020), and imprecision with denominator data on traveller numbers for Australia (eg, some travellers were moved between states on domestic flights which is not captured in the official data we used). Additionally, case numbers are constantly changing, due to the number of reclassifications caused by false positives and duplications. We also did not assess the change in the quarantine system failures over time due to the relatively small number of failure events. The risk of system failures is probably highly dynamic on a month-by-month basis as traveller volumes, infection rates, and quarantine processes change, and as vaccination rates among border workers and in the wider community increase.

To substantially reduce the risk of SARS-CoV-2 incursion out of quarantine (until such time as enough of the population is vaccinated), the most obvious action is to reduce arrivals, or even suspend arrivals, from high infection locations (as New Zealand and Australia temporarily did for travel from India and other high risk countries in April 2021^22^). Beyond this, there are a range of other potential improvements in ongoing arrangements and processes as detailed in Table 2. Furthermore, the start of quarantine-free travel between Australia and New Zealand (also known as a “green zone”) in April 2021 provides an opportunity to benchmark COVID-19 border control policies and practices, identify potential improvements in both countries, and harmonise best practices across the region. The green zone further intertwines the biosecurity status of both nations and it is therefore even more important to lower the risk of border failures that could disrupt such travel. This shift from a one-size-fits-all strategy to a risk-based approach to border management can be summarised as a ‘traffic light’ approach.^23^

**Table 2:** List of potential policy and operational options for improved COVID-19 control associated with quarantine systems in Australia and New Zealand, including measures to reduce the numbers of infected people arriving into quarantine facilities.

| **Policy option** | **Description** | **Our priority rankings** |
| --- | --- | --- |
| 1. Cap travel from high prevalence countries and/or suspend for a period | Reduce the in-flow of travellers by reducing or suspending flights to Australia and NZ<sup>a</sup> from very high incidence countries where the pandemic is out of control. These governments have the legal powers to put conditions on the existing rights of their citizens to enter their country of citizenship (ie, on public health grounds). | Top priority |
| 2. Pre-departure testing plus/minus pre-departure quarantine | Expand existing requirements for pre-departure testing to additional traveller source countries. Pre-departure testing could be expanded from not only a PCR<sup>b</sup> test within 72 hours of departure to also add a rapid test at the airport immediately before departure (given many infected may have started shedding the virus in the previous 72 hours and most, but not all, of such cases will be detected by a rapid test even though it has lower sensitivity). Of note is that such arrangements are considered legally acceptable (see the above row).<br>Pre-departure quarantine (eg, for a week), would provide additional assurance, but this would probably need to be in a transport hub (eg, at an airport hotel at Singapore or Hawaii) where NZ and Australian officials were permitted access to ensure quality processes. Even if establishing a formal facility is shown to be impractical, all incoming travellers could be asked to self-quarantine as strictly as possible in the week before travel, eg, by a request through the passenger booking system (see Policy Option 4). | Top priority |
| 3. Pre-departure vaccination | Make travel contingent on completing a course of approved vaccination. This measure assumes the vaccine is at least partially effective at preventing transmission. This requirement needs further investigation and development. | Uncertain |
| 4. Use passenger booking systems to reduce infection risk | Require passengers to declare pre-departure COVID-19<sup>c</sup> precautions via the system that they use to book spaces in quarantine facilities prior to travel. Such a system is operating in NZ and could be adopted more widely in Australia. | High priority |
| 5. Increase in-flight precautions | Explore means to reduce the risk of in-flight infection as documented on a flight to NZ.<sup>25</sup> This could be via more stringent enforcement of mask wearing in airports and on flights, and the use of higher-efficacy masks (although fit can be critical to the level of protection offered by some masks and respirators) and/or double masking.<br>An experimental study conducted by researchers at the US CDC<sup>d</sup> found that a medical procedure mask alone blocked 56.1% of particles from a simulated cough, and a cloth mask alone blocked 51.4%, while the combination of a cloth mask covering a medical procedure mask (double masking) blocked 85.4% of cough particles.<sup>26</sup> In a second experiment, double masking of the source dummy (which produced simulated cough particles) reduced the cumulative exposure of an unmasked receiver dummy by 82.2%, and when the source was unmasked and the receiver was fitted with a double mask, the receiver's cumulative exposure was reduced by 83.0%.<sup>26</sup> When both the source and the receiver were double masked, the cumulative exposure of the receiver was reduced by 96.4%.<sup>26</sup><br>Another laboratory-based study that used humans rather than dummies to compare the FFE<sup>e</sup> of commonly available masks worn singly, doubled, or in combination found that on average, across all masks and volunteers, adding a second medical procedure mask improved mean FFE from 55% when single masking to 66% when | High priority |

| Policy option | Description | Our priority rankings |
| --- | --- | --- |
|  | double masking.<sup>27</sup> Specifically, wearing a procedure mask under a cloth face covering produced the largest improvement in overall FFE (from 66% to 81%).<sup>7</sup> The improvement in FFE was likely due to the reduction of leaks between the mask and skin.<sup>27</sup> While the findings of these studies may not be representative of double masking in real world settings, they suggest the potential for both improved source control (ie, reduced exposure from infected wearers) and reduced wearer exposure (ie, reduced exposure of uninfected wearers). Minimizing talking when masks are displaced during eating and drinking, and improved ventilation and spacing requirements on flights might also be worthwhile. |  |
| 6. Reduce infection risk at airports and transit hubs | Ensure measures are in place at departure airports and transit hubs to minimize the risk of cross infection (eg, through physical distancing and mask use). | Medium priority |
| 7. Improve local transport arrangements | Ensure sufficient physical distancing of travellers on arrival and in transit to quarantine (eg, lowering density on buses). For such arrangements, higher efficacy masks or double masking could be required (see "5. Increase in in-flight precautions" above). | Medium priority |
| 8. Shift to discrete quarantine units | Shift some or all quarantine facilities to rural military bases or camps where discrete units (eg, mobile homes or caravans) could be appropriately spatially separated. The success (to date – see Table A1 in the Appendix) of the Howard Springs facility (a converted workers' camp<sup>24</sup>) should be considered. This approach allows for natural ventilation and eliminates shared indoor spaces. If spaces were limited, then these settings could be used for travellers from the highest risk countries. | High priority |
| 9. Restrict hotel quarantine in large cities to low-risk travellers | Reserve hotel quarantine in large cities to the lowest risk category of travellers, with hotels in more minor cities being used for the highest risk category of travellers. However, the risk/benefit analysis of such changes would need to consider airport access and if the additional travelling to minor cities poses excessive additional risk. | High priority |
| 10. Expand use of PCR testing of saliva in facility workers (and travellers) | Expand the regular (daily) use of PCR testing of saliva of facility workers to all facilities in both countries. This approach could also be considered for all travellers, albeit potentially still combined with existing testing regimens. In view of increased transmissibility of new variants, consideration should be given to testing of all workers in border-associated occupations (eg, providing airline meals and laundry services) at least twice per week. Documented negative tests at appropriate frequency should be an occupational requirement for all border workers instead of the self-report systems (as currently used in NZ and Australia). | High priority |
| 11. Accelerate or mandate vaccination for quarantine staff | Vaccinate all quarantine workers against COVID-19 and redeploy all unvaccinated workers from the front line. This measure will be particularly valuable when vaccines are known to prevent transmission in addition to protecting recipients from illness. | As of April 2021, this measure was nearing completion in some jurisdictions |
| 12. Cohorting complete flights of travellers | Cohorting of flights means that all returnees arriving into a country go to the same quarantine facility until that facility is full and then the intake switches to another facility and so on. This approach is designed to reduce cross-infection in such facilities. This system was introduced in NZ on 16 May 2021. | Medium priority |
| 13. Upgrade processes at quarantine facilities | Upgrade processes at quarantine facilities in terms of eliminating shared spaces (eg, no shared exercise areas and shared smoking areas), in particular ensuring that day cohorts do not mix under any circumstances. Ventilation improvements could also be considered with limiting the use of rooms to only those with external windows or a balcony. | Medium priority |
| 14. Prosecute rule breaking in quarantine facilities | Enforce quarantine facility rules more rigorously. Rule breaking, which is relatively common in NZ facilities,<sup>28</sup> could start to be prosecuted (given no prosecutions during 2020). | Medium priority |
| 15. Improve conditions for quarantine staff | Improve working conditions for the staff in quarantine facilities to minimize the risk of overwork (which may increase the risk of PPE failures) or of workers taking on other part-time jobs in other settings. For example, in February 2021 there were still concerns by NZ health workers about staffing inadequacies in these facilities. Some states in Australia have banned front-line quarantine staff from working second jobs. | High priority |
| 16. Improve management of travellers who smoke | Introduce specific measures for travellers who are nicotine dependent to reduce their need to smoke in designated areas during their travel and while in managed quarantine (eg, nicotine replacement treatment as a requirement for travel). | Medium priority |
| 17. Add post-quarantine control measures | Introduce a post-quarantine period of home-quarantine to reduce the risk of local transmission arising from undetected infections in people leaving hotel quarantine facilities (which may arise from either exceptionally long incubation periods or cross infection during quarantine stays). Post-quarantine testing could also be used to detect such infections. | Medium priority |
| 18. Mandate the use of digital contact tracing tools | Mandate quarantine workers to use digital technologies (eg, the Bluetooth function on the COVID Tracer smartphone apps) to facilitate contact tracing in the event of a border failure. Travellers could be required to use such technologies for two weeks after completing their time in quarantine. There is also a case for travellers using these tools within quarantine as (at least in NZ) quarantine facilities are sometimes evacuated for fire alarms and burst water pipes. | Medium priority |

<sup>a</sup> NZ: New Zealand
<sup>b</sup> PCR: polymerase chain reaction
<sup>c</sup> COVID-19: coronavirus disease 2019
<sup>d</sup> US CDC: United States Centers for Disease Control and Prevention
<sup>e</sup> FFE: fitted filtration efficiency

## Conclusions

In summary, Australia and New Zealand have had 32 COVID-19 identified failures arising from quarantine systems up to 15 June 2021. Quarantine system failures can be costly in terms of lives and economic impacts such as lockdowns. Ongoing improvements or alternatives to hotel-based quarantine are required.

## Data Availability

The datasets analyzed during the current study were derived from public resources available from the New Zealand Ministry of Business Innovation & Employment, the New Zealand Ministry of Health. the Australia Bureau of Statistics. and the Australian Department of Health.

https://www.mbie.govt.nz/business-and-employment/economic-development/covid-19-data-resources/managed-isolation-and-quarantine-data/

https://www.health.govt.nz/our-work/diseases-and-conditions/covid-19-novel-coronavirus/covid-19-data-and-statistics/covid-19-case-demographics

https://www.abs.gov.au/statistics/industry/tourism-and-transport/overseas-travel-statistics-provisional/latest-release

https://www.health.gov.au/resources/collections/coronavirus-covid-19-common-operating-picture

## Appendix

**Appendix Table A1:** List of COVID-19 border control failures associated with quarantine systems in Australia and New Zealand during 2020 and up to 15 June 2021.

| Event | Extent of known spread | Details |
| --- | --- | --- |
| **Australia** |  |  |
| Rydges Hotel – Victoria "second wave" outbreak (late May to late October 2020) | Over 19,800 cases,<sup>29, 30</sup> and over 800 deaths<sup>31</sup> | Genomic testing indicated that 99% of Victoria's second wave of community COVID-19<sup>a</sup> cases were linked to transmission events related to returned travellers infecting workers at the Rydges Hotel in Carlton and the Stamford Plaza Hotel (see row below) in Melbourne's CBD<sup>b</sup>, which were used as facilities for quarantine.<sup>32</sup> Specifically, around 90% of cases can be traced back to a single family of four that returned to Australia in mid-May and were quarantined at the Rydges Hotel.<sup>33</sup> The virus then spread from the infected workers to the community, with high rates of local transmission.<sup>32</sup> The outbreak led to a stringent lockdown for 112 days in the state, with particularly strict measures in the major city of Melbourne.<sup>34</sup> At least nine people employed in Melbourne's hotel quarantine program tested positive between late July and early October 2020, although the cases may have been a reflection of substantial community transmission in Melbourne at the time rather than additional hotel quarantine failures.<sup>35</sup> Two of those cases worked while infectious.<sup>35</sup> |
| Stamford Plaza Hotel - Victoria "second wave" outbreak (late May to late October 2020) | See row above | Almost 10% of cases in Victoria's "second wave" outbreak were attributable to an outbreak at the Stamford Hotel in mid-June.<sup>32</sup> The outbreak was traced back to international travellers who returned to Australia in early June,<sup>32</sup> and then was spread by security guards who worked at the facility.<sup>36</sup> |
| Marriot Hotel at Circular Quay in Sydney, NSW<sup>h</sup> (August 2020) | 2 security guards | A security guard at the Marriot Hotel at Circular Quay in Sydney tested positive for COVID-19 on 15 August.<sup>37</sup> Genomic sequencing linked the infection to a returned overseas traveller at the facility.<sup>37</sup> A second security guard subsequently tested positive.<sup>38</sup> |
| Parafield outbreak in South Australia (December 2020) | 33 cases<sup>31</sup> | Genomic testing indicated that Adelaide's Parafield cluster was linked to transmission events related to a returned traveller in a quarantine hotel infecting workers in the facility, possibly due to poor ventilation at the facility.<sup>32</sup> The virus spread from the workers to the community, resulting in a strict lockdown.<sup>32</sup> |
| Quarantine hotel facility cleaner infected in Sydney, NSW (December 2020) | A single worker | A quarantine hotel worker (a cleaner) who completed shifts at two quarantine hotels, the Ibis Hotel and the Novotel in Darling Harbour in Sydney, tested positive in early December.<sup>39</sup> There was also spread in a facility with 5 returned travellers being infected. |
| Airport Bus Driver NSW (December 2020) | A single worker | A Sydney van driver who drives international flight crew to and from Sydney Airport, tested positive for COVID-19 on 17 December 2021.<sup>40</sup> Although he was symptomatic and worked for 3 days before being tested, this did not cause an outbreak and it was contained to one case.<sup>40</sup> |
| Avalon outbreak in NSW (January 2021) | 151 cases (as of 11 January 2021)<sup>41</sup> | It has been reported that genomic sequencing suggests that the strain is of US origin and entered Australia via an infectious returned traveller who entered hotel quarantine upon arrival.<sup>39</sup> It is currently unknown how the virus spread to the community and source investigations were still underway (as per January 2021). Case numbers include those from the linked Croydon and Wollongong clusters, but not the linked Black Rock cluster in Victoria (27 cases as of 5 January 2021) due to separate reporting by states. |
| Berala outbreak in NSW (January 2021) | 26 cases (as of 11 January 2021)<sup>41</sup> | It has been reported that genomic sequencing linked the outbreak back to infectious international travellers.<sup>42</sup> A patient transport worker transferred infectious travellers from Sydney Airport to a hospital.<sup>41, 42</sup> Another patient transport worker who was a close contact of the first then also tested positive.<sup>41, 42</sup> The virus then spread from the second infected worker into the community, seeding the Berala outbreak.<sup>42</sup> |
| Hotel Grand Chancellor outbreak in Brisbane, Queensland (January 2021) | 6 cases (as of 13 January 2021)<sup>43</sup> | A hotel quarantine worker in Brisbane tested positive in early January 2021 for the Alpha variant of COVID-19.<sup>44</sup> This case led to a three-day lockdown in the greater Brisbane area, while contact tracers worked to ensure there was no community transmission of the strain.<sup>45</sup> It was later revealed that there were six genomically linked cases within the quarantine facility, including the hotel quarantine worker and five returned travellers.<sup>43</sup> |
| Four Points by Sheraton hotel, Western Australia (January 2021) | A single worker | A Perth quarantine hotel security guard, who worked a second job as a ride share driver, tested positive to the UK strain of SARS-CoV-2<sup>c</sup>, sparking a five-day lockdown for 80 per cent of the state's population, in an attempt to stop any further transmission.<sup>46</sup> It's believed he was exposed to the virus on the 26 January when he worked at Four Points by Sheraton, on the same floor where a quarantined returned traveller with a confirmed case of infection with the Alpha variant, was staying. After feeling sick on 28 January, the case visited a GP<sup>d</sup>, and tested positive on 30 January. The exact cause is unknown, but it's believed poor ventilation could be a factor.<sup>47</sup> This comes a week after Western Australia committed to daily testing of the hotel quarantine staff, instead of weekly.<sup>48</sup> Quarantine hotel staff, including cleaners, security guards and catering staff, are no longer allowed to have second jobs but will receive a pay increase of about 40 per cent as compensation.<sup>46</sup> It has been announced an inquiry will be held.<sup>47</sup> |
| Grand Hyatt Hotel, Victoria (February 2021) | A single worker | One 26-year-old staff member at the Grand Hyatt Hotel (an isolation facility that was used for players and support staff for the Australian Open) tested positive for a highly transmissible variant of COVID-19 (ie, the B.1.1.7 or Alpha variant) in February 2021, leading to heightened restrictions.<sup>49</sup> The close contacts of the case all tested negative, and the outbreak did not spread into the wider community.<sup>49</sup> |
| Holiday Inn Hotel Cluster, Victoria (February 2021) | 22 cases | Twenty-two cases have been linked to the Holiday Inn cluster in Melbourne, where an infected returned traveller used a nebuliser, causing an outbreak and a 5-day lockdown.<sup>50</sup> There were no further cases detected in the community during the lockdown as all confirmed cases had already been informed of their status as close contacts and were in isolation for the duration of their infectious period. Cases included returned travellers, hotel quarantine staff and their families, as well as a Melbourne Airport worker.<sup>50</sup> |
| Princess Alexandra Hospital Doctor outbreak, Brisbane, Queensland (March 2021) | 6 cases<sup>51</sup> | A doctor and a nurse (see row below) who worked at the Princess Alexandra Hospital while unvaccinated were linked to two separate COVID-19 outbreaks.<sup>51</sup> While both outbreaks were the highly transmissible Alpha variant of the disease and originated at the same hospital, they were from two different sources.<sup>51</sup> The doctor tested positive on 12 March 2021, with subsequent infections in 5 other persons.<sup>51</sup> These outbreaks resulted in a 3-day lockdown just before the Easter holiday.<sup>51</sup> |
| Princess Alexandra Hospital Nurse outbreak, Brisbane, Queensland (March 2021) | 13 cases<sup>52</sup> | A nurse (see row above) who worked on a COVID-19 ward at the Princess Alexandra Hospital while unvaccinated tested positive in late March.<sup>51</sup> Genome sequencing links the case to an overseas traveller from India who was being cared for at the hospital.<sup>53</sup> The nurse's sister, a number of attendees at a "hen's party", and several other contacts were subsequently infected.<sup>51</sup> |
| Sofitel Wentworth in Sydney, NSW (March 2021) | A single security guard | A security guard at the Sofitel Wentworth in Sydney tested positive for COVID-19 on 13 March through routine surveillance testing.<sup>54</sup> Genomic sequencing linked the case to a returned traveller and subsequent testing revealed that another returned traveller was also infected within the facility.<sup>54</sup> |
| Mercure Hotel Cluster in Perth, Western Australia (April 2021) | 3 cases | A mother and her four-year-old daughter who had returned from the UK, and a traveller who had returned from China contracted the virus while in hotel quarantine.<sup>55</sup> The mother and child's infections were detected on 16 April and genomically linked to a couple who had returned from India on April 10 and subsequently tested positive for the Alpha variant.<sup>55</sup> All of |
|  |  | the travellers were staying in adjacent rooms on the same hotel floor.<sup>55</sup> The returned traveller from China tested negative on release from hotel quarantine in Perth on 17 April, but later tested positive for COVID-19 in Melbourne.<sup>56</sup> The case then spent up to five days in the community in Perth while infectious, and two linked community cases were identified.<sup>57</sup> The outbreak led to a 3 day lockdown in Perth and the neighbouring Peel region,<sup>56</sup> and a brief suspension in quarantine-free travel between Western Australia and NZ.<sup>57</sup> The ventilation at the Mercure hotel had previously been identified as the riskiest among Western Australia's 10 quarantine hotels, with an engineer finding that the corridors had no independent airflow with oxygen supply leaking from the adjoining rooms.<sup>55</sup> Chief Health Officer Andy Robertson had recommended that the Mercure Hotel no longer operate as a quarantine facility on 14 April.<sup>55</sup> |
| Community cases in NSW (May 2021) | 2 known cases (possibly more) | On 5 May 2021, a man in his 50s from Sydney was diagnosed with COVID-19.<sup>58</sup> His wife subsequently tested positive.<sup>58</sup> The cases were genomically linked to a traveller who returned from the US and was moved to a quarantine facility on 28 April.<sup>59</sup> However, an epidemiological link between the traveller and the cases has yet to be established,<sup>58</sup> suggesting that there may be additional cases in the community. Additionally, fragments of coronavirus were detected in wastewater samples in Sydney.<sup>59</sup> The cases did not result in a lockdown in NSW, but additional restrictions, including compulsory masking and limits on indoor gatherings were put in place.<sup>58</sup> Additionally, the situation led the NZ Government to temporarily pause their quarantine-free travel arrangement with NSW for 48 hours, and a close contact of one of the cases was placed in a MIQ facility in Christchurch after travelling to NZ.<sup>58</sup> |
| Pan Pacific Hotel in Perth, Western Australia (May 2021) | 3 cases | A security guard at the Pan Pacific Hotel in Perth tested positive on 1 May.<sup>60</sup> He worked on the same floor as two infected travellers (one from the US and one from Indonesia).<sup>60</sup> He had had the first Pfizer vaccination, but not the second.<sup>60</sup> Two of the seven people that he lived with subsequently tested positive.<sup>60</sup> This was an event for which Western Australia was still subject to restrictions from a previous outbreak.<sup>60</sup> |
| Whittlesea Community Outbreak in Victoria, stemming from the Playford Hotel in South Australia (May 2021) | At least 63 cases (outbreak was still active as of 23 June 2021) | A traveller who returned from India via the Maldives and Singapore completed the required stay in quarantine in South Australia.<sup>61</sup> After leaving quarantine on 4 May, he then travelled directly to his home in Victoria. He subsequently developed symptoms and tested positive.<sup>61</sup> It is thought that the traveller likely contracted the virus while in quarantine in South Australia; it has been reported that the traveller stayed next door to another person who tested positive for the virus.<sup>61</sup> This later caused a widespread outbreak, leaking into aged care, and resulted in a 14-day lockdown followed by tight restrictions.<sup>62</sup> |
| West Melbourne Delta Variant Cluster, Victoria (May 2021) | 15 known cases (possibly more) | This outbreak has been genomically linked to a hotel quarantine case that arrived in Victoria from Sri Lanka on 8 May 2021 and tested positive on the same day.<sup>63</sup> The case was subsequently moved from the Novotel Ibis quarantine hotel in Melbourne's CBD to the Holiday Inn on 9 May, and then was released from quarantine on 23 May 2021.<sup>63</sup> The variant was then detected in members of a family in Victoria that were known to have travelled to parts of NSW, and had the same genomic sequence as the hotel quarantine case.<sup>63</sup> However, there is still uncertainty about the link between these cases, as no formal epidemiological link has been established, suggesting that there may have been additional cases in the community.<sup>63</sup> |
| Airport Limo Driver in NSW (June 2021) | 1 case (outbreak was still active as of 23 June 2021) | A 60-year-old airport limo driver, who was involved in transporting international flight crew to and from Sydney Airport, tested positive for COVID-19 on 15 June.<sup>64</sup> It's believed he transported a crew of three from a FedEx freight plane before he became infectious, on 11 June.<sup>64</sup> This is currently being investigated, as authorities believe this man breached NSW quarantine and testing protocols.<sup>64</sup> |
| **New Zealand** |  |  |
| Auckland outbreak (August 2020) | A total of 179 cases, with 3 deaths<sup>65</sup> | The cause of this outbreak remains unknown, but genomic work probably provides the best evidence to this being a border facility (isolation or quarantine facility) failure: "There are a large number of similar genomes which are from the UK<sup>e</sup>, which would seem to suggest the UK is the most likely source of any unknown importation".<sup>66</sup> This was at a time when 40% of cases in NZ quarantine/isolation facilities did not have genomic work on the virus infecting them (ie, there was not enough complete virus in the samples). It was also estimated<sup>66</sup> that there was only a very tiny risk of this outbreak being a continuation of the March/April spread of the pandemic in NZ: "Our Bayesian phylogenetic analysis ... estimates that there is a 0.4% probability that case 20VR2563 is in the "sister clade" of the Auckland cluster." Finally, the chance of the outbreak being from contaminated imported food was also considered very unlikely: "Our Bayesian phylogenetic analysis ... shows that the estimated mutation rate on the branch leading to the cluster is not a lot smaller than elsewhere in the tree, lending little weight to the possibility that the virus lay dormant on packing material for a long period of time." |
| Border facility maintenance worker infected (August 2020) | A single worker | A shared lift environment in a quarantine hotel (the Rydges Hotel in Central Auckland) was the source suspected by officials,<sup>67</sup> with the sharing being only minutes apart.<sup>66</sup> The genomic sequencing indicated the same virus infecting the worker as per a recent traveller in the same facility.<sup>66</sup> |
| Border facility health worker infected (September 2020) | A single worker | This was a work-related infection associated with a hotel facility used for isolating infected cases (the Jet Park Hotel, Auckland), given that the case was linked via genomic sequencing to 3 cases within the facility.<sup>68</sup> Some details of the full investigation report were provided to the media and these suggested that while the nurse wore PPE<sup>f</sup>, there was a period where the associated patient did not have a mask on during treatment.<sup>69</sup> This situation could have contributed to a failure of the PPE worn by the nurse. |
| Traveller-related outbreak / Crowne Plaza, Christchurch border facility (September 2020) | The 2 returnees and 4 others | Three returnees (Cases A, B, and C) tested positive for SARS-CoV-2 while in an MIQ facility (ie, these cases were caught at the border).<sup>70</sup> Evidence suggests that two returnees (Cases D and E) were then infected through suspended aerosol particles while in the same hotel quarantine facility (Cases D and E were staying in the room next to Case C), before then moving into the community.<sup>70, 71</sup> These infected returnees appear to have then infected another person (Case G), potentially on a charter flight after leaving the facility.<sup>70, 71</sup> A household contact of Cases D and E was also infected (Case F).<sup>70, 72</sup> Two household contacts of Case G were also infected.<sup>70</sup> |
| Border facility health worker (Case A) in Christchurch (November 2020) | A single worker | This was a work-related infection associated with a facility used for isolating infected cases. Both this case (and "Case B" below) had the virus genome sequencing linked to infection in a group of international mariners in the same hotel facility but with different virus subtypes in each case.<sup>67</sup> "The finding supports the current theory that there were two separate events infecting both workers at the facility." These cases of infected health workers appear to have contributed to border control nurses threatening strike action if they were not supplied with improved PPE. One estimate was that 12 of the mariners were infected on arrival in NZ, but with subsequent spread within the facility a total of 31 mariners were ultimately infected.<sup>73</sup> |
| Another border facility health worker (Case B) (see above) | A single worker | This was a separate work-related infection associated with a border control facility – see in the row above. |
| Defence Force worker outbreak (November 2020) | The worker, a co-worker and 4 others (total of 6 cases) | This was a work-related infection in a Defence Force worker associated with a hotel border facility in Auckland (used for isolating known infected cases). "The genome sequencing we have conducted on Case A's test result shows a direct link to two travellers who are part of a family group in the quarantine facility".<sup>74</sup> The route of transmission to one of the |
|  |  | community cases remains a mystery (albeit they worked in the same locality within Auckland City). Associated with these cases, the Prime Minister made statements around the need for further risk reduction.<sup>75</sup> |
| Traveller infectious after leaving a quarantine facility (January 2021) | 1 traveller | A traveller was identified as being infectious in the community after leaving a quarantine facility (Pullman, Auckland). The traveller reportedly had the Beta variant (lineage B.1.351) of the pandemic virus.<sup>76</sup> Genome sequencing has linked this case to another traveller who was in the same facility.<sup>77</sup> The transmission mechanism has not been precisely identified with investigations still proceeding as of mid-February 2021. However, officials consider it likely to have been a separate transmission event to the one in the following row. |
| Travelers infectious after leaving a quarantine facility (January 2021) | 2 travellers and a contact | Two travellers (a parent and child) were identified as infectious in the community after being infected with the Beta variant of the pandemic virus, with a link to a quarantine facility (Pullman, Auckland).<sup>78</sup> A close contact (the mother of the child) also became infected.<sup>79</sup> The transmission mechanism has not been precisely identified with investigations still proceeding as of mid-February 2021. |
| MIQ<sup>9</sup> workers (March 2021) | 3 workers | A cleaner at the Grand Millennium Hotel MIQ facility in Auckland (Case A) tested positive during routine surveillance testing on 22 March 2021.<sup>80</sup> While a household contact returned a weak positive the next day,<sup>81</sup> subsequent tests were negative.<sup>82</sup> The MIQ worker had recently been vaccinated (with the Pfizer/BioNTech vaccine) on 23 February, with a second dose on 16 March,<sup>81</sup> although they tested positive before full protection from the vaccine could be expected. Their family had not yet been vaccinated.<sup>81</sup> Genome sequencing revealed that Case A was infected with a highly transmissible variant (Alpha) and was linked to a traveller in the facility who had arrived on 13 March.<sup>83</sup> In early April, two unvaccinated security guards at the same facility tested positive (Cases B and C) and genome sequencing linked the two cases to Case A.<sup>84, 85</sup> |

<sup>a</sup> COVID-19: coronavirus disease 2019
<sup>b</sup> CBD: central business district
<sup>c</sup> SARS-CoV-2: severe acute respiratory syndrome coronavirus 2
<sup>d</sup> GP: general practitioner
<sup>e</sup> UK: United Kingdom
<sup>f</sup> PPE: personal protective equipment
<sup>g</sup> MIQ: managed isolation and quarantine
<sup>h</sup> NSW: New South Wales

## References

1. Baker M, Wilson N, Blakely T. Elimination may be the optimal response strategy for covid-19 and other emerging pandemic diseases. BMJ 2020; 371: m4907. doi: 4910.1136/bmj.m4907.

2. World Health Organization. Considerations for quarantine of contacts of COVID-19 cases: Interim guidance, 2020. https://www.who.int/publications/i/item/considerations-for-quarantine-of-individuals-in-the-context-of-containment-for-coronavirus-disease-(covid-19) (accessed Aug 2021)

3. Ritchie H, Ortiz-Ospina E, Beltekian D, et al. New Zealand: Coronavirus pandemic country profile published online at OurWorldInData.org 2021 (updated Apr 2021). https://ourworldindata.org/coronavirus/country/new-zealand?country=NZL~AUS (accessed Apr 2021).

4. New Zealand Ministry of Health. COVID-19: Vaccine data, 2021 (updated Apr 2021). https://www.health.govt.nz/our-work/diseases-and-conditions/covid-19-novel-coronavirus/covid-19-data-and-statistics/covid-19-vaccine-data (accessed Apr 2021).

5. Ilanbey S, Dexter R, Estcourt D. All Victorian frontline hotel quarantine workers to be vaccinated by Thursday. The Age (Victoria/Coronavirus pandemic) 2021; 5 Apr. https://www.theage.com.au/national/victoria/no-new-local-covid-19-cases-as-calls-grow-for-mass-vaccination-centres-to-be-fast-tracked-20210405-p57ghc.html (accessed Apr 2021).

6. Ministry of Business Innovation & Employment. Managed isolation and quarantine data. New Zealand, 2021. https://www.mbie.govt.nz/business-and-employment/economic-development/covid-19-data-resources/managed-isolation-and-quarantine-data/ (accessed Jun 2021).

7. Ministry of Health. COVID-19: Case demographics 2021 (updated Jun 2021). https://www.health.govt.nz/our-work/diseases-and-conditions/covid-19-novel-coronavirus/covid-19-data-and-statistics/covid-19-case-demographics (accessed Jun 2021).

8. Australian Bureau of Statistics. Overseas Travel Statistics, Provisional (Released 16 June 2021). https://www.abs.gov.au/statistics/industry/tourism-and-transport/overseas-travel-statistics-provisional/latest-release (accessed Jun 2021).

9. Australian Department of Health. Coronavirus (COVID-19) common operating picture, 2021. https://www.health.gov.au/resources/collections/coronavirus-covid-19-common-operating-picture (accessed Jun 2021).

10. O’Brien J, Monteiro C, Rezny M, et al. COVID-19 in Australia, 2021 (updated Apr 2021). https://www.theage.com.au/national/victoria/three-new-covid-19-cases-linked-to-holiday-inn-cluster-emerge-20210219-p573wq.html (accessed Apr 2021).

11. Baden LR, El Sahly HM, Essink B, et al. Efficacy and safety of the mRNA-1273 SARS-CoV-2 vaccine. N Engl J Med 2021; 384: 403–416.

12. Brouwer PJM, Brinkkemper M, Maisonnasse P, et al. Two-component spike nanoparticle vaccine protects macaques from SARS-CoV-2 infection. Cell 2021; 184: 1188-1200,e1119.

13. Haas EJ, Angulo FJ, McLaughlin JM, et al. Impact and effectiveness of mRNA BNT162b2 vaccine against SARS-CoV-2 infections and COVID-19 cases, hospitalisations, and deaths following a nationwide vaccination campaign in Israel: an observational study using national surveillance data. Lancet 2021; 397:1819-1829.

14. Geoghegan JL, Ren X, Storey M, et al. Genomic epidemiology reveals transmission patterns and dynamics of SARS-CoV-2 in Aotearoa New Zealand. Nat Commun 2020; 11: 6351–6357.

15. COVID research updates: 7 January — Evidence grows of a new coronavirus variant’s swift spread. Nature 2021, 8 Jan. https://www.nature.com/articles/d41586-020-00502-w (accessed Apr 2021).

16. Voysey M, Costa Clemens S, Madhi SA, et al. Single dose administration and the influence of the timing of the booster dose on immunogenicity and efficacy of ChAdOx1 nCoV-19 (AZD1222) vaccine: a pooled analysis of four randomised trials. Lancet 2021, 397: 881–891.

17. Emary KRW, Golubchik T, Aley PK, et al. Efficacy of ChAdOx1 nCoV-19 (AZD1222) vaccine against SARS-CoV-2 variant of concern 202012/01 (B.1.1.7): an exploratory analysis of a randomised controlled trial. Lancet 2021; 397: 1351–1362.

18. Covid-19: Why Australia’s quarantine rules are changing. Radio New Zealand (World) 2021; 9 Jan. https://www.rnz.co.nz/news/world/434278/covid-19-why-australia-s-quarantine-rules-are-changing (accessed Apr 2021).

19. Morrah M. Revealed: The strict new coronavirus rules for New Zealand’s COVID-19 Defence Force staff. Newshub 2020; 25 Nov. https://www.newshub.co.nz/home/new-zealand/2020/11/revealed-the-strict-new-coronavirus-rules-for-new-zealand-s-covid-19-defence-force-staff.html (accessed Apr 2021).

20. Premier of Victoria. A stronger quarantine program to protect what we’ve built [press release]. 2020; 30 Nov. https://www.premier.vic.gov.au/stronger-quarantine-program-protect-what-weve-built (accessed Apr 2021).

21. The Victorian government reaches a deal with the Commonwealth over new quarantine hub. ABC News 2021; 4 Jun. https://www.abc.net.au/news/2021-06-04/new-quarantine-hub-to-be-built-in-victoria/100189846 (accessed Jun 2021).

22. Cheng D. Watch: Covid Response Minister Chris Hipkins reveals travel ban for ‘very high risk countries’. NZ Herald 2021; 23 Apr. https://www.nzherald.co.nz/nz/watch-covid-response-minister-chris-hipkins-reveals-travel-ban-for-very-high-risk-countries/ZRZAJIGZ6Q3OBWPX6C2ZB7BLPM/ (accessed Apr 2021).

23. Wilson N, Baker M. New Zealand needs a ‘traffic light’ system to stop COVID-19 creeping in at the border. The Conversation 2020; 4 Nov. https://theconversation.com/new-zealand-needs-a-traffic-light-system-to-stop-covid-19-creeping-in-at-the-border-149262 (accessed Apr 2021).

24. Northern Territory Government. Mandatory supervised quarantine for repatriated Australians 2020. https://coronavirus.nt.gov.au/travel/quarantine/mandatory-supervised-quarantine-for-repatriated-australians (accessed Apr 2021).

25. Swadi T, Geoghegan JL, Devine T, et al. Genomic evidence of in-flight transmission of SARS-CoV-2 despite predeparture testing. Emerg Infect Dis 2021; 27: 687–693.

26. Brooks JT, Beezhold DH, Noti JD, et al. Maximizing fit for cloth and medical procedure masks to improve performance and reduce SARS-CoV-2 transmission and exposure. MMWR Morb Mortal Wkly Rep 2021; 70: 254–257.

27. Sickbert-Bennett EE, Samet JM, Prince SE, et al. Fitted filtration efficiency of double masking during the COVID-19 pandemic. JAMA Intern Med 2021; Apr 16: e212033.

28. Cheng D. Covid 19 coronavirus: Chafing under the rules - 76 bubble breaches in four months. New Zealand Herald (Politics) 2020; 28 Nov. https://www.nzherald.co.nz/nz/covid-19-coronavirus-chafing-under-the-rules-76-bubble-breaches-in-four-months/5WLODXO7S4SBCZY7UWBWKSZOUI/ (accessed Apr 2021).

29. Zhang L, Tao Y, Zhuang G, et al. Characteristics analysis and implications on the COVID-19 reopening of Victoria, Australia. Innovation (N Y) 2020; 1: 100049.

30. Department of Health and Human Services, State Government of Victoria. Tracking coronavirus in Victoria, 2020. https://www.dhhs.vic.gov.au/tracking-coronavirus-victoria (accessed Apr 2021).

31. Government of South Australia. COVID-19 update 22 December 2020 [press release], 2020. https://www.sahealth.sa.gov.au/wps/wcm/connect/public+content/sa+health+internet/about+us/news+and+media/all+media+releases/covid-19+update+22+december+2020 (accessed Apr 2021).

32. Board of Inquiry into the COVID-19 Hotel Quarantine Program. COVID-19 hotel quarantine inquiry final report and recommendations, Volume 1. Victorian Government Printer, 2020.

33. Taylor J. Hotel quarantine linked to 99% of Victoria’s covid cases, inquiry told. The Guardian (Victoria) 2020; 18 Aug. https://www.theguardian.com/australia-news/2020/aug/18/hotel-quarantine-linked-to-99-of-victorias-covid-cases-inquiry-told (accessed Apr 2021).

34. Mercer P. Covid: Melbourne’s hard-won success after a marathon lockdown. BBC News (Australia) 2020; 26 Oct. www.bbc.com/news/world-australia-54654646 (accessed Apr 2021).

35. Covid-19 coronavirus: Officials still searching for the source of Sydney outbreak. NZ Herald (World) 2020; 20 Dec. https://www.nzherald.co.nz/world/covid-19-coronavirus-officials-still-searching-for-source-of-sydney-outbreak/UCDN5SOFNOKFM232YTXG4AWDK4/ (accessed Apr 2021).

36. Lucas C. Stamford hotel blames government and guards for outbreak. The Age (Politics/Victoria/Coronavirus pandemic) 2020; 2 Jul. https://www.theage.com.au/politics/victoria/stamford-hotel-blames-government-and-guards-for-outbreak-20200702-p558e6.html (accessed Apr 2021).

37. NSW coronavirus investigation after Sydney hotel quarantine security guard tests positive. ABC News 2020; 18 Aug. https://www.abc.net.au/news/2020-08-18/nsw-coronavirus-hotel-quarantine-security-guard-investigated/12570336 (accessed Apr 2021).

38. Aubusson K, Chung L. Australian health authorities racing to contain cluster as NSW records 14 new covid-19 cases. Stuff 2020; 29 Aug. https://www.stuff.co.nz/world/australia/300094865/australian-health-authorities-racing-to-contain-cluster-as-nsw-records-14-new-covid19-cases (accessed Apr 2021).

39. Tsirtsakis A. Is it time for an national approach to hotel quarantine? newsGP (Clinical) 2020; 4 Dec. https://www1.racgp.org.au/newsgp/clinical/is-it-time-for-a-national-approach-to-hotel-quaran (accessed Apr 2021).

40. Nguyen K. NSW coronavirus warning as Sydney airport driver tests positive for COVID-19. ABC News 2020; 16 Dec. https://www.abc.net.au/news/2020-12-16/nsw-confirms-new-locally-acquired-coronavirus-case/12988866 (accessed Jun 2021).

41. NSW Health. Covid-19 weekly surveillance in NSW: Epidemiological week 52, ending 26 December 2020. NSW Government; 2021. https://www.health.nsw.gov.au/Infectious/covid-19/Documents/covid-19-surveillance-report-20201226.pdf (accessed Apr 2021).

42. Lathouris O. Berala cluster: Syney COVID outbreak linked to BWS where two infected staff worked throughout Christmas. Nine News (National) 2021; 4 Jan. https://www.9news.com.au/national/coronavirus-nsw-update-berala-bws-cluster-not-linked-to-avalon-sydney-northern-beaches-covid19-strain-genomic-testing-finds/26e4bfd0-df34-42d6-852c-5ac774bc8a11 (accessed Apr 2021).

43. Incident response set up following confirmation of Brisbane hotel cluster [press release]. Queensland Government, 2021; 13 Jan. https://www.health.qld.gov.au/news-events/doh-media-releases/releases/incident-response-set-up-following-confirmation-of-brisbane-hotel-cluster (accessed Apr 2021).

44. Gramenz E. Quarantine hotel worker tests positive to coronavirus in Brisbane, Queensland records two new cases. ABC News 2021; 7 Jan. https://www.abc.net.au/news/2021-01-07/coronavirus-queensland-quarantine-hotel-worker-tests-positive/13034046 (accessed Apr 2021).

45. Queensland Government. Greater Brisbane 3-day lockdown 2021 https://www.qld.gov.au/health/conditions/health-alerts/coronavirus-covid-19/current-status/greater-brisbane-lockdown (accessed Apr 2021).

46. Manfield E. WA coronavirus hotel quarantine security companes agree to ban on second jobs. ABC News 2021; 10 Feb. https://www.abc.net.au/news/2021-02-10/wa-hotel-quarantine-security-guards-to-end-second-jobs-next-week/13139172 (accessed Apr 2021).

47. Hendrie D. Latest hotel quarantine leak proves ‘we’re not learning’. newsGP. 2021; 1 Feb. https://www1.racgp.org.au/newsgp/clinical/latest-hotel-quarantine-leak-proves-we-re-not-lear (accessed Apr 2021).

48. Woodley M. National Cabinet strengthens Australia’s border controls. newsGP 2021; 8 Jan. https://www1.racgp.org.au/newsgp/clinical/national-cabinet-strengthens-australia-s-border-co (accessed Apr 2021).

49. No new local cases in Victoria, amid hotal quarantine leak. The New Daily 2021; 5 Feb. https://thenewdaily.com.au/news/coronavirus/2021/02/05/hotel-quarantine-victoria-zero/ (accessed Apr 2021).

50. Estcourt D, Sakkal P. One in intensive care as Holiday Inn COVID-19 cluster grows to 22. The Age (Victoria/Coronavirus pandemic) 2021; 19 Feb. Available from: https://www.theage.com.au/national/victoria/three-new-covid-19-cases-linked-to-holiday-inn-cluster-emerge-20210219-p573wq.html (accessed Apr 2021).

51. Motherwell S. How two clusters from one hospital triggered the Brisbane lockdown. ABC News 2021; 30 Mar. https://www.abc.net.au/news/2021-03-30/brisbane-lockdown-clusters-coronavirus-explained-pa-hospital/100037608 (accessed Apr 2021).

52. Historic case linked to Brisbane COVID-19 cluster [press release]. Queensland Health, 2021; 11 Apr. https://www.health.qld.gov.au/news-events/doh-media-releases/releases/historic-case-linked-to-brisbane-covid-19-cluster (accessed Apr 2021).

53. Brisbane COVID-19 clusters explained [press release]. Queensland Health, 2021; 30 Mar. https://www.health.qld.gov.au/news-events/doh-media-releases/releases/brisbane-covid-19-clusters-explained (accessed Apr 2021).

54. Kontominas B. NSW quarantine hotel worker tests positive to coronavirus. ABC News 2021; 14 Mar. https://www.abc.net.au/news/2021-03-14/nsw-records-one-new-covid-19-case-in-hotel-quarantine-worker/13246856 (accessed Apr 2021).

55. McNeill H, de Kruijff P. Perth plunged into three-day lockdown over Mercure Hotel cluster. WA Today (Coronavirus pandemic) 2021; 23 Apr. https://www.watoday.com.au/national/western-australia/perth-plunged-into-lockdown-over-mercure-hotel-cluster-20210423-p57lwb.html (accessed Apr 2021).

56. Perth lockdown: WA premier wants designated quarantine facilities. Radio New Zealand (World/COVID-19) 2021; 24 Apr. https://www.rnz.co.nz/news/world/441176/perth-lockdown-wa-premier-wants-designated-quarantine-facilities (accessed Apr 2021).

57. Covid-19: ‘Low’ risk to NZ from Perth case - Health Ministry. Radio New Zealand (New Zealand/COVID-19) 2021; 24 April. https://www.rnz.co.nz/news/national/441154/covid-19-low-risk-to-nz-from-perth-case-health-ministry (accessed Apr 2021).

58. NSW records no new COVID-19 cases in community, ahead of NZ travel bubble decision. 1 News 2021; 8 May. https://www.tvnz.co.nz/one-news/world/nsw-records-no-new-covid-19-cases-in-community-ahead-nz-travel-bubble-decision (accessed Jun 2021).

59. Cheng D. Covid 19 coronavirus: Thousands of travellers’ plans disrupted as flights from NSW paused for 48 hours. NZ Herald (Politics) 202; 7 May. https://www.nzherald.co.nz/nz/covid-19-coronavirus-thousands-of-travellers-plans-disrupted-as-flights-from-nsw-paused-for-48-hours/VHG4T2FY5IUZVPND7GYZJMALVE/ (accessed Jun 2021).

60. De Poloni G. Perth hotel quarantine guard tests positive for COVID-19, along with two others. ABC News 2021; 1 May. https://www.abc.net.au/news/2021-05-01/perth-hotel-quarantine-guard-tests-positive-for-covid-19/100109788 (accessed Jun 2021).

61. Victoria records COVID-19 case at Wollert, north of Melbourne, after man leaves SA hotel quarantine. ABC News 2021; 11 May. https://www.abc.net.au/news/2021-05-11/victoria-covid-19-case-man-north-of-melbourne/100131038 (accessed Jun 2021).

62. Lockdown restrictions in regional Victoria to ease as Melbourne COVID outbreak grows to ABC News 2021; 3 Jun. https://www.abc.net.au/news/2021-06-03/new-covid-cases-detected-in-victoria-as-lockdown-continues/100186750 (accessed Jun 2021).

63. Dunstan J. How the Delta COVID variant likely jumped Victorian hotel quarantine and started Melbourne outbreak. ABC News 2021; 8 Jun. https://www.abc.net.au/news/2021-06-08/melbourne-covid-outbreak-delta-strain-link-hotel-quarantine/100183468 (accessed Jun 2021).

64. Malone U. How the potentially ‘inexcusable’ actions fo a limo driver put Sydney on COVID-19 alert. ABC News 2021; 18 Jun. https://www.abc.net.au/news/2021-06-17/nsw-quarantine-worker-may-have-breached-health-order/100223120 (accessed Jun 2021).

65. Ministry of Health. COVID-19: Source of cases 2021 [updated 23 Jun 2021]. https://www.health.govt.nz/our-work/diseases-and-conditions/covid-19-novel-coronavirus/covid-19-data-and-statistics/covid-19-source-cases (accessed Jun 2021).

66. Hadfield J, Douglas J, Geoghegan J, et al. Re-emergence of community transmission in Aotearoa New Zealand -Genomic overview of the Auckland Outbreak. Institute of Environmental Science and Research (ESR) 2021. https://nextstrain.org/community/narratives/ESR-NZ/GenomicsNarrativeSARSCoV2/2020-10-01?n=5 (accessed Oct 2020).

67. 1 case of COVID-19 in quarantine worker [press release]. New Zealand Ministry of Health, 2020; 7 Nov. https://www.health.govt.nz/news-media/media-releases/1-case-covid-19-quarantine-worker (accessed Apr 2021).

68. 1 new case of COVID-19 [press release]. New Zealand Ministry of Health, 2020; 14 Sep. https://www.health.govt.nz/news-media/media-releases/1-new-case-covid-19-18 (accessed Apr 2021).

69. Quinn R. Covid-infected nurse ‘did everything right’, report finds. Radio New Zealand 2020; 6 Nov. https://www.rnz.co.nz/news/national/429973/covid-infected-nurse-did-everything-right-report-finds (accessed Apr 2021).

70. Eichler N, Thornley C, Swadi T, et al. Transmission of Severe Acute Respiratory Syndrome Coronavirus 2 during border quarantine and air travel, New Zealand (Aotearoa). Emerg Infect Dis 2021; 27: 1274–1278.

71. No new cases of COVID-19 [press release]. New Zealand Ministry of Health, 2020; 2 Oct. https://www.health.govt.nz/news-media/media-releases/no-new-cases-covid-19-50 (accessed Apr 2021).

72. 2 new cases of COVID-19 [press release]. New Zealand Ministry of Health, 2020; 27 Sep. https://www.health.govt.nz/news-media/media-releases/2-new-cases-covid-19-21 (accessed Apr 2021).

73. Canterbury District Health Board. International mariners quarantine: Summary of official information request 2021 (updated 22 Jan 2021). https://www.cdhb.health.nz/about-us/document-library/international-mariners-quarantine/ (accessed Apr 2021).

74. 4 new cases of COVID-19 in managed isolation [press release]. New Zealand Ministry of Health, 2020; 9 Nov. https://www.health.govt.nz/news-media/media-releases/4-new-cases-covid-19-managed-isolation (accessed Apr 2021).

75. Cheng D. Covid 19 coronavirus: Jacinda Ardern’s immediate moves to minimise risk for MIQ workers. New Zealand Herald (Politics) 2020; 9 Nov. https://www.nzherald.co.nz/nz/politics/covid-19-coronavirus-jacinda-arderns-immediate-moves-to-minimise-risk-for-miq-workers/ZGJEQB77N6Z3JIFUY3KMJJSETU/ (accessed Apr 2021).

76. 2 cases of COVID-19 in managed isolation; update on Northland case [press release]. New Zealand Ministry of Health, 2020; 26 Jan. https://www.health.govt.nz/news-media/media-releases/2-cases-covid-19-managed-isolation-update-northland-case (accessed Apr 2021).

77. Update on Northland case, and 6 cases of COVID-19 in managed isolation [press release]. New Zealand Ministry of Health, 2021; 25 Jan. https://www.health.govt.nz/news-media/media-releases/update-northland-case-and-6-cases-covid-19-managed-isolation (accessed Apr 2021).

78. 3 new cases of COVID-19 at the border and an update on border-related cases in Auckland [press release]. New Zealand Ministry of Health, 2021; 28 Jan. https://www.health.govt.nz/news-media/media-releases/3-new-cases-covid-19-border-and-update-border-related-cases-auckland (accessed Apr 2021).

79. 7 new cases of COVID-19 [press release]. New Zealand Ministry of Health, 2021; 4 Feb. https://www.health.govt.nz/news-media/media-releases/7-cases-covid-19 (accessed Apr 2021).

80. Ministry reporting positive COVID-19 case at the border [press release]. New Zealand Ministry of Health, 2021; 22 Mar. https://www.health.govt.nz/news-media/media-releases/ministry-reporting-postive-covid-19-case-border (accessed Apr 2021).

81. Covid-19: MIQ worker’s family returns three negative, one weak positive result - Hipkins. Radio New Zealand (COVID-19) 2021; 23 Mar. https://www.rnz.co.nz/news/covid-19/438962/covid-19-miq-worker-s-family-returns-three-negative-one-weak-positive-result-hipkins (accessed Apr 2021).

82. No new community cases; 3 cases of COVID-19 in managed isolation [press release]. New Zealand Ministry of Health, 2021; 26 Mar. https://www.health.govt.nz/news-media/media-releases/no-new-community-cases-3-cases-covid-19-managed-isolation-0 (accessed Apr 2021).

83. One new border-related case; 19 cases of COVID-19 in managed isolation; 4 historical cases [press release]. New Zealand Ministry of Health, 2021; 8 Apr. https://www.health.govt.nz/news-media/media-releases/one-new-border-related-case-19-cases-covid-19-managed-isolation-4-historical-cases (accessed Apr 2021).

84. No new community cases; 7 cases of COVID-19 in managed isolation; 1 historical case [press release]. New Zealand Ministry of Health, 2021, 12 Apr. https://www.health.govt.nz/news-media/media-releases/no-new-community-cases-7-cases-covid-19-managed-isolation-1-historical-case (accessed Apr 2021).

85. New border-related positive COVID-19 case [press release]. New Zealand Ministry of Health, 2021; 11 Apr. https://www.health.govt.nz/news-media/media-releases/new-border-related-positive-covid-19-case (accessed Apr 2021).

